# Immune profiling of COVID-19 vaccine responses in people with multiple sclerosis on B cell-depleting therapy

**DOI:** 10.1101/2023.12.04.23299409

**Authors:** Griffith B. Perkins, Christopher M. Hope, Cheng Sheng Chai, Matthew J. Tunbridge, Sebastian Sterling, Kevin Webb, Joey Yap, Arthur Eng Lip Yeow, Makutiro G. Masavuli, Svjetlana Kireta, James D. Zuiani, Anouschka Akerman, Anupriya Aggarwal, Vanessa Milogiannakis, Matthew B. Roberts, William Wilson, Plinio R. Hurtado, Stuart Turville, Branka Grubor-Bauk, Simon C. Barry, P. Toby Coates, Janakan Ravindran, Pravin Hissaria

**Affiliations:** Central and Northern Adelaide Renal and Transplantation Service, Royal Adelaide Hospital; Adelaide, Australia 5000; Adelaide Medical School, University of Adelaide; Adelaide, Australia 5000; Immunology Directorate, SA Pathology; Adelaide, Australia 5000; Molecular Immunology, Robinson Research Institute, University of Adelaide; Adelaide, Australia 5000; Department of Paediatric Medicine, Women’s and Children’s Hospital; North Adelaide, Australia 5000; Central Adelaide Local Health Network; Adelaide, Australia 5000; Department of Neurology, Royal Adelaide Hospital; Adelaide, Australia 5000; Viral Immunology Group, Basil Hetzel Institute for Translational Health Research, University of Adelaide; Woodville, Australia 5011; Kirby Institute, University of New South Wales; Sydney, Australia 2052; Infectious Diseases, Royal Adelaide Hospital; Adelaide, Australia 5000; Northern Adelaide Local Health Network; Elizabeth Vale, Australia 5112; Department of Immunology, Royal Adelaide Hospital; Adelaide, Australia 5000

## Abstract

**Background and Objective:** People with multiple sclerosis (pwMS) receiving B cell-depleting therapies have impaired antibody responses to vaccination. In a proportion of individuals, repeat vaccination against COVID-19 leads to seroconversion. We sought to describe the immune phenotype of pwMS on ocrelizumab, and identify clinical and immunological determinants of an effective vaccine response.

**Methods:** This was a single-centre, prospective cohort study. Peripheral blood samples were collected from pwMS receiving ocrelizumab (n = 38) pre and post administration of a third dose of mRNA COVID-19 vaccine. Immunogenicity was measured by T cell IFNγ ELISpot, antibody titres, and live virus neutralisation. Humoral immunity was benchmarked against pwMS receiving natalizumab (n = 15), and against a correlate of real-world protection (50% reduction in incidence of infection) from SARS-CoV-2 ancestral and omicron BA.5 variants. The peripheral immune phenotype was comprehensively assessed by flow cytometry, and potential clinical and phenotypic determinants of response to vaccination identified.

**Results:** Immune cell populations relevant to disease and vaccine response were altered in pwMS receiving ocrelizumab versus natalizumab treatment, including depleted CD20-expressing B cell, T cell and NK cell populations, and elevated CD27^+^CD38^+^ T cell and ‘NK8’ cell frequencies. Following a third vaccine dose, 51% of pwMS on ocrelizumab were seropositive for SARS-CoV-2 receptor-binding-domain IgG, and 25% and 14% met the threshold for effective neutralisation of live SARS-CoV-2 ancestral and omicron BA.5 virus, respectively. B cell frequency at the time of vaccination, but not time since ocrelizumab infusion, was positively correlated with antibody response, while a strong negative correlation was observed between CD56^bright^ NK cell frequency and antibody response in the ocrelizumab group. In this exploratory cohort, CD3^−^CD20^+^ B cells (% of lymphocytes; OR=3.92) and CD56^bright^ NK cells (% of NK cells; OR=0.94) were predictive of an effective neutralising antibody response in second dose non-responders (AUC: 0.98).

**Discussion:** Ocrelizumab treatment was associated with an altered immune phenotype, including recently described T cell and NK populations with potential roles in disease pathogenesis. However, seroconversion was severely impaired by ocrelizumab, and less than half of those who seroconverted following a third vaccine dose demonstrated effective immunity against SARS-CoV-2 ancestral or omicron BA.5. B cell frequency was associated with an effective antibody response, while immunomodulatory CD56^bright^ NK cells were identified as a potential negative determinant of response in those with inadequate B cell numbers. Immune phenotype rather than time since ocrelizumab infusion may help to stratify individuals for prophylaxis.

## INTRODUCTION

Disease modifying therapies (DMTs) employed in the treatment of relapsing remitting multiple sclerosis (RRMS) can affect the immune response to vaccination. Common DMTs include the B cell-depleting anti-CD20 monoclonal antibody, ocrelizumab (Ocrevus), and the anti-α4-integrin monoclonal antibody, natalizumab (Tysabri). B cell depleting therapies are associated with increased risk of severe COVID-19 disease ^1–5^ and impaired humoral immune response to vaccination ^6–10^ in pwMS and other patient populations. In contrast, natalizumab is non-depleting, and no impairment in vaccine response has been identified for pwMS receiving treatment with natalizumab.^6, 7, 11^

Recent studies have identified seroconversion rates by pwMS on B cell-depleting therapies as ∼50% following three COVID-19 vaccine doses, amongst the lowest of any group, with antibody titres in those who do seroconvert inferior to healthy individuals.^5–8, 10, 12, 13^ As such, it has been difficult to interpret the level of protection afforded these patients, particularly against antibody-escape variants such as omicron BA.5, and for clinicians and policy makers to make informed decisions with respect to booster dosing and existing and emerging prophylactic measures.

Here we describe cellular and humoral immune responses to a third dose of mRNA-platform (BNT162b2 Pfizer or mRNA1273 Moderna) COVID-19 vaccine in pwMS receiving ocrelizumab. Seroconversion rates were compared against pwMS receiving natalizumab, and against a correlate of real-world protection from infection based on live virus neutralisation of ancestral and omicron BA.5 in order to estimate the level of protection afforded by a third vaccine dose. Finally, we describe phenotypic changes in the peripheral immune system associated with ocrelizumab use, and identify potential clinical and phenotypic predictors of response to vaccination.

## RESULTS

|  | All<br>Participants<br>(N = 53) | Ocrelizumab<br>(N = 38) | Natalizumab<br>(N = 15) |
| --- | --- | --- | --- |
| Sex |  |  |  |
| Female, N (%) | 37 (69.8) | 26 (68.4) | 11 (73.3) |
| Male, N (%) | 16 (30.2) | 12 (31.6) | 4 (26.7) |
| Age, median (IQR), y | 44 (35-54) | 44 (36-53) | 44 (33-54) |
| MS type |  |  |  |
| Relapsing-remitting (%) | 53 (100) | 38 (100) | 15 (100) |
| Progressive (%) | 0 (0) | 0 (0) | 0 (0) |
| Time from last ocrelizumab infusion to 3rd dose median (IQR), d | N/A (N/A) | 158 (113-183) | N/A (N/A) |
| First Vaccine, N (%) |  |  |  |
| BNT162b2 (Pfizer) | 50 (93.3) | 35 (92.1) | 15 (100) |
| mRNA-1273 (Moderna) | 1 (1.9) | 1 (2.6) | 0 (0) |
| ChAdOx1 (AstraZeneca) | 2 (3.8) | 2 (5.3) | 0 (0) |
| Second Vaccine, N (%) |  |  |  |
| BNT162b2 (Pfizer) | 51 (96.2) | 36 (94.7) | 15 (100) |
| mRNA-1273 (Moderna) | 1 (1.9) | 1 (2.6) | 0 (0) |
| ChAdOx1 (AstraZeneca) | 1 (1.9) | 1 (2.6) | 0 (0) |
| Third Vaccine, N (%) |  |  |  |
| BNT162b2 (Pfizer) | 51 (96.2) | 38 (100) | 13 (86.7) |
| mRNA-1273 (Moderna) | 2 (3.8) | 0 (0) | 2 (13.3) |
| ChAdOx1 (AstraZeneca) | 0 (0) | 0 (0) | 0 (0) |
| Time between third dose and sample median (IQR), d | 29 (28-155) | 28 (28-31) | 166 (49-191) |
| Contracted COVID prior to study, N (%) | 9 (17.0) | 4 (10.5) | 5 (33.3) |
| Contracted COVID during follow-up, N (%) | 16 (30.2) | 11 (28.9) | 5 (41.7) |
| Remdesivir, N (%)* | 2 (12.5) | 2 (18.2) | 0 (0) |
| Sotrovimab, N (%)* | 3 (18.8) | 3 (27.3) | 0 (0) |
| Nirmatrelvir/ritonavir, N (%)* | 2 (12.5) | 2 (18.2) | 0 (0) |
| Tixagevimab/cilgavimab, N (%)* | 7 (43.8) | 7 (63.6) | 0 (0) |

\* % of COVID positive during follow-up

Study demographics are presented in **Table 1**. Thirty-eight participants receiving ocrelizumab were enrolled in the study and peripheral blood samples were collected immediately prior to, and four weeks after, the third vaccine dose. For comparison of immune phenotype and vaccine response, samples were collected from 15 pwMS receiving natalizumab at a single time point post third dose. Median age (44 vs 44 years), sex (68% vs 73% female) and primary COVID-19 vaccine type (95% vs 100% mRNA-platform) were similar between treatment groups, and all participants received an mRNA vaccine (BNT162b2 Pfizer or mRNA1273 Moderna) as a third dose. The median time from vaccination to blood sample collection was 28 days (IQR: 28–31) for the ocrelizumab group, and 166 days (IQR: 49–191) for the natalizumab group. Five participants in the natalizumab group, and four in the ocrelizumab group, had COVID-19 prior to the study and were excluded from analyses unless indicated.

### Booster vaccination improves SARS-CoV-2-specific antibody and T cell responses

Seroconversion was assessed with the ‘Elecsys Anti-SARS-CoV-2 S’ assay (Roche, Basel, Switzerland), through clinical diagnostic services. Following the third vaccine dose, a proportion of pwMS on ocrelizumab with detectable anti-receptor binding domain (RBD-) IgG increased from 29.7% to 51.5% (Figure 1A). A concordant increase was observed in the median titre of anti-Spike (S-) IgG (in-house ELISA; AUC 7.149 vs 160.5, *p* = 0.0096), but not S-IgM nor S-IgA (Figure 1B). Third vaccine dose improved T cell responses in 24/34 individuals (71%), with the median frequency of Spike-specific, IFNγ-secreting T cells increasing from 1046 to 1903 spot-forming units (SFU) per 10^6^ cells (*p* < 0.0001, Figure 1C).

**Figure 1.**
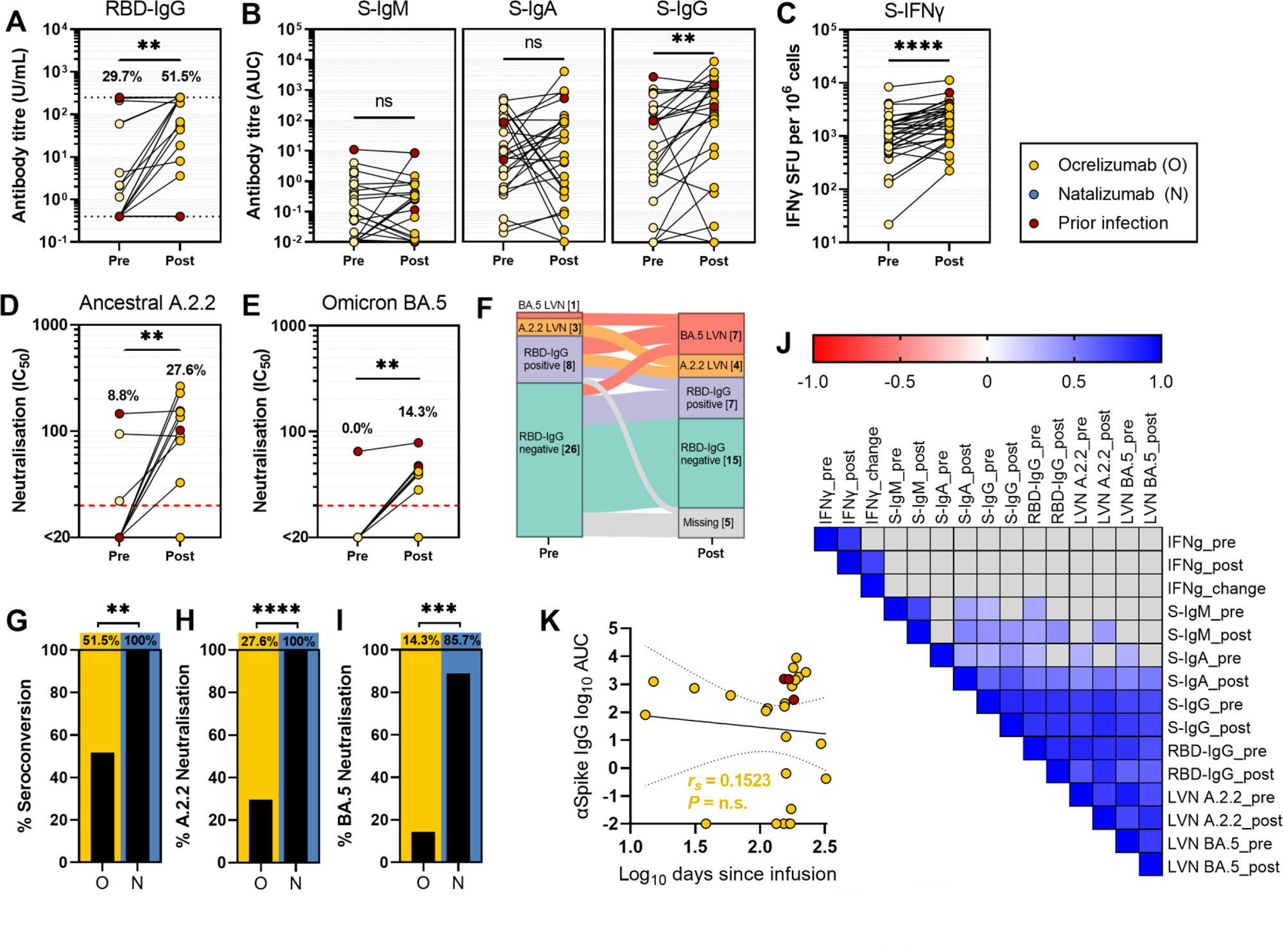
Effect of a third vaccine dose on protective immunity to SARS-CoV-2 ancestral and omicron BA.5 variants in pwMS on B cell depletion therapy. **(A)** Anti-SARS-CoV-2 receptor binding domain IgG (RBD-IgG) titres. Dashed lines demarcate the upper (250 U/mL) and lower (0.4 U/mL) detection limits of the assay, and percentage seropositivity (>0.4 U/mL) pre and post booster vaccination is indicated. **(B)** Anti-SARS-CoV-2 Spike IgM (S-IgM), IgA (S-IgA) and IgG (S-IgG) titres measured as area under the curve (AUC). **(C)** SARS-CoV-2 Spike-specific, IFNγ-secreting T cells (S-IFNγ) presented as IFNγ spot-forming units per 10^6^ peripheral blood mononuclear cells (PBMCs). **(D-E)** Serological neutralisation of live SARS-CoV-2 virus ancestral A.2.2 (D) and omicron BA.5 (E) strains, pre and post booster vaccination. Red dashed lines indicate the target threshold for effective neutralisation correlating with 50% protection from infection. Percentage effective neutralisation is indicated. **(F)** Overview of changes in sero-status with a third vaccine dose. Participants are classified by maximum response achieved, where BA.5 LVN > A.2.2 LVN > RBD-IgG positive > RBD-IgG negative. **(G-I)** Comparison of humoral immunity following a third vaccine dose in pwMS receiving ocrelizumab versus natalizumab, presented graphically and numerically. Treatment groups are compared by percentage of participants achieving seroconversion of anti-RBD IgG (H), and effective neutralisation of ancestral A.2.2 (I) and omicron BA.5 (J) variants. (J) Spearman’s correlation analysis of vaccine response parameters pre and post third vaccine dose. **(K)** Relationship between anti-Spike IgG post-third vaccination versus days between last ocrelizumab infusion and third vaccine dose administration with Spearman’s correlation analysis. **Statistical significance by Wilcoxon-matched pairs signed rank test (straight bars) or two-sided Fisher’s exact test (crooked bars). Participants with prior SARS-CoV-2 infection (red) were not included in the statistics presented, nor in statistical significance calculations. ****P<0.0001; ***P<0.001; **P<0.01; *P<0.05; ns, non-significant.** RBD-IgG, anti-SARS-CoV-2 receptor binding domain immunoglobulin-G; S-IgM/A/G, anti-SARS-CoV-2 Spike protein immunoglobulin; A.2.2/BA.5-LVN, SARS-CoV-2 ancestral/omicron BA.5 live virus neutralisation; S-IFNγ, SARS-CoV-2 Spike protein-specific IFNγ ELISpot.

Anti-nucleocapsid (NC-) IgG, which is routinely used for assessment of past infection, was below detection in all pwMS on ocrelizumab, including in four individuals with previous PCR positive SARS-CoV-2 infections (data not shown). The five participants in the natalizumab group with documented prior infection all returned positive NC-IgG titres.

### Neutralising antibody responses are inadequate in pwMS receiving ocrelizumab

The capacity of serum to neutralise live SARS-CoV-2 virus correlates closely with real-world protection from infection and disease.^14^ To estimate vaccine effectiveness, Khoury *et al* defined the relationship between live virus neutralisation titre and risk of infection with SARS-CoV-2, and derived a value that correlates with 50% protection from infection (20.2% of the mean neutralising titre of first-wave convalescent individuals; 95% CI 14.4% to 28.4%). To establish an equivalent threshold in the present study, serum samples from 20 first-wave convalescent individuals were analysed side-by-side with the study samples, and a target threshold for effective neutralisation of IC_50_ = 20 defined (see Methods). Effective neutralisation of ancestral SARS-CoV-2 increased from 8.8% to 27.6% with a third vaccine dose in the ocrelizumab cohort (Figure 1D). Fewer patients demonstrated effective neutralisation of the omicron BA.5 variant, increasing from 0.0% to 14.3% with the third dose (Figure 1E).

Antibody responses following the third dose were compared to participants treated with the non-depleting monoclonal antibody natalizumab. Relative to the natalizumab cohort, pwMS receiving ocrelizumab demonstrated reduced seroconversion of RBD-IgG (51.5% vs 100%, *p* = 0.0067; Figure 1H), and significantly lower rates of effective neutralisation of SARS-CoV-2 ancestral (27.6% vs 100%, *p* < 0.0001; Figure 1I) and omicron BA.5 (14.3% vs 80.0%, *p* = 0.0004; Figure 1J) variants, despite a longer median time from vaccination to sample collection for the natalizumab group (median: 28 vs 166 days).

During the follow-up period, 7/34 infection naïve participants (21%) from the ocrelizumab group and 2/10 infection naïve participants (20%) from the natalizumab group reported contracting COVID-19. No cases resulted in hospitalisation. All patients in the ocrelizumab group and none in the natalizumab group, underwent treatment with antivirals.

### Time since ocrelizumab infusion is not associated with vaccine response

Clinical and demographic parameters were assessed for association with antibody responses to the third vaccine dose in the ocrelizumab group. In a multivariate linear regression model, only male sex was associated with antibody response (anti-RBD IgG: *β* = 131.2, 95% CI 33.99 to 228.3, *p* = 0.0104; A.2.2 neutralisation: *β* = 73.93, 95% CI 2.745 to 145.1, *p* = 0.0425). Strong positive correlations were observed between pre and post vaccination SARS-CoV-2-specific immune measures (Figure 1J), however pre-existing immunity (i.e. antibody and T cell responses resulting from the primary vaccination schedule) was not associated with the magnitude of T cell or antibody response to the third dose. Notably, time between last ocrelizumab infusion and vaccination did not predict, nor correlate with, S-IgG response to vaccination (r_s_ = 0.1523, *p* = 0.2277; Figure 1K).

### Immune phenotype associated with ocrelizumab versus natalizumab treatment

Severe impairment in protective immunity associated with ocrelizumab treatment, and the lack of a relationship between modifiable clinical parameters and vaccine responses, prompted us to evaluate the effect of ocrelizumab on immune phenotype to identify immune determinants of vaccine response. One-hundred-and-one immune phenotypes were assessed in the peripheral blood of pwMS receiving ocrelizumab and natalizumab (Supplementary Figure 1-3). Principle component analysis based on these phenotype variables resulted in clustering of patients by treatment, supporting the notion that treatment is the major determinant of immune variation within the cohort (Figure 2A). Eight immune parameters were increased, and 14 decreased, in the ocrelizumab group compared with the natalizumab group, based on a desired false discovery rate of 1% for significance (Figure 2B). As expected, pwMS on ocrelizumab demonstrated reductions in CD19^+^, CD20^+^ and CD19^+^CD20^+^ B cells as a proportion of lymphocytes (Figure 2B-E). To visualise differences in the phenotype of B cells between treatment groups, a two-dimensional plot of concatenated immune phenotype data was constructed using the t-distributed stochastic neighbour embedding (t-SNE) dimensionality reduction algorithm (Figure 2C). B cell subpopulation clusters were defined by FlowSOM, and relative frequencies of B cell populations visualised for each treatment group (Figure 2C). The B cell compartment in the ocrelizumab cohort was enriched for immature transitional B cells, reflecting a step-wise reconstitution of the B cell compartment following depletion (Figure 2C).

**Figure 2.**
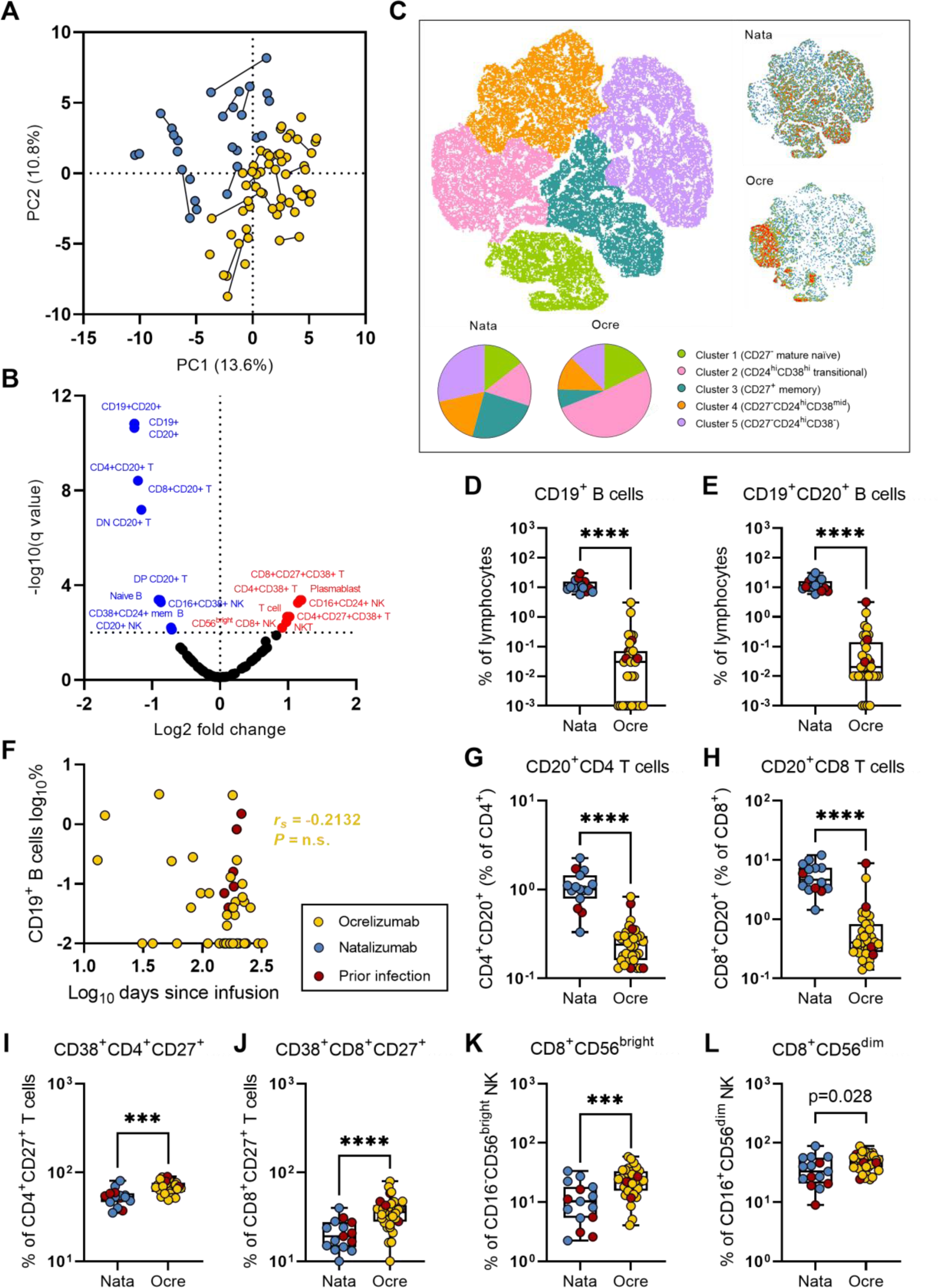
Immune phenotype associated with B cell depletion therapy. **(A)** Principle component analysis (PCA) of 101 peripheral blood immune phenotype parameters analysed in pwMS on ocrelizumab or natalizumab. PC1 and PC2 explain 13.6% and 10.8% of variation in immune phenotype, respectively. Paired pre and post vaccination samples are joined with a line. **(B)** Volcano plot identifying significant differences (false discovery rate < 1%) in immune phenotype parameters between the ocrelizumab and natalizumab treatment groups. **(C)** Visualisation of B cell subset bias in pwMS receiving ocrelizumab versus natalizumab. The t-distributed stochastic neighbour embedding (tSNE) dimensionality reduction algorithm was applied to concatenated phenotype data for a representative cohort (equal number of ocrelizumab and natalizumab samples and CD19^+^ events), and broad B cell subpopulations defined and coloured by FlowSOM clustering (left). Relative frequencies of B cell subpopulations in representative samples from ocrelizumab and natalizumab cohorts are visualised by density plot of equal event number and quantified with pie charts (right). **(D-E)** Comparison of CD19^+^ (D) and CD19^+^CD20^+^ (E) B cell frequencies as a percentage of lymphocytes between treatment groups at the time of vaccination. **(F)** Relationship of days since last ocrelizumab infusion and CD19^+^ B cells (% of lymphocytes) with Spearman’s correlation analysis. **(G-L)** Comparison of selected immune phenotype parameters between treatment groups immediately prior to vaccination. T cells were defined as CD3^+^CD19^−^CD14^−^ CD56^+^ cells, and NK cells as CD3^−^CD19^−^CD14^−^CD56^+^/CD16^+^ cells, within the physical lymphocyte gate. Frequency of CD20^+^CD4^+^ (G) and CD20^+^CD8^+^ (H) T cells assessed as a percentage of lymphocytes. Percentage of CD4^+^CD27^+^ (I) and CD8^+^CD27^+^ (J) T cells expressing the activation marker CD38. **(K-L)** Percentage of CD56^bright^CD16^−^ (K) and CD56^dim^CD16^+^ (L) NK cells positive for CD8. **Differences between treatment groups identified by Mann-Whitney with two-stage step-up method to correct for multiple comparisons. P-values for presented comparisons (D-E, G-L) are unadjusted: ****P<0.0001; ***P<0.001; *P<0.05. Participants with prior SARS-CoV-2 infection (red) were not included in statistical significance calculations.**

In addition to ablation of B cells, proportions of CD20-expressing populations within the CD4^+^, CD8^+^, double positive (CD4^+^CD8^+^) and double negative (CD4^−^CD8^−^) T cell compartments were significantly reduced (Figure 2B, G-H). CD20^+^CD4^+^ and CD20^+^CD8^+^ T cells, which have been implicated in the pathogenesis of MS,^15–17^ represented the greatest difference between groups, with a 4.2-fold reduction in the proportion of CD4^+^ T cells (geometric mean: 1.0% vs 0.24%, *p* < 0.0001) and 9.7-fold reduction in the proportion of CD8^+^ T cells (4.9% vs 0.50%, *p* < 0.0001) expressing CD20 in the ocrelizumab group (Figure 2G-H).

Conversely, the proportion of CD4^+^ T cells expressing the co-stimulatory receptor CD27, recently proposed as a mechanism underlying the efficacy of B cell depletion therapy in MS,^18^ was elevated in the ocrelizumab group (87% vs 93%, *p* = 0.0209). Expression of the activation marker CD38 was increased in both CD4^+^CD27^+^ (52% vs 68%, *p* < 0.0001) and CD8^+^CD27^+^ (19% vs 34%, *p* < 0.0001) T cells in the ocrelizumab group (Figure 2I-J).

Finally, CD8^+^ NK (‘NK8’) cells were recently identified as an immunomodulatory cell type associated with reduced risk of relapse in pwMS not receiving treatment.^19^ CD8^+^ NK cells were elevated in the ocrelizumab group as a percentage of both CD56^bright^CD16^−^ immunomodulatory (9.2% vs 21.3%, *p* < 0.0001) and CD56^dim^CD16^+^ conventional NK cells (32% vs 46%, *p =* 0.0284; Figure 2K-L).

### B cells and CD56^bright^ NK cells are associated with vaccine response in pwMS receiving ocrelizumab

Correlation analysis was performed to describe the relationship between immune phenotype parameters and vaccine response in pwMS receiving ocrelizumab (Figure 3A). Strong positive correlations were observed between B cell populations and post-vaccination humoral immune responses in the ocrelizumab cohort (Figure 3A). B cell (CD20^+^CD19^+^) frequency at the time of vaccination correlated strongly with anti-Spike and anti-RBD IgG titres post-vaccination, and with serum neutralisation of ancestral and omicron BA.5 SARS-CoV-2 variants, in the ocrelizumab group (Figure 2B). Similar trends were observed in the natalizumab cohort, which did not reach significance (Figure 2B).

**Figure 3.**
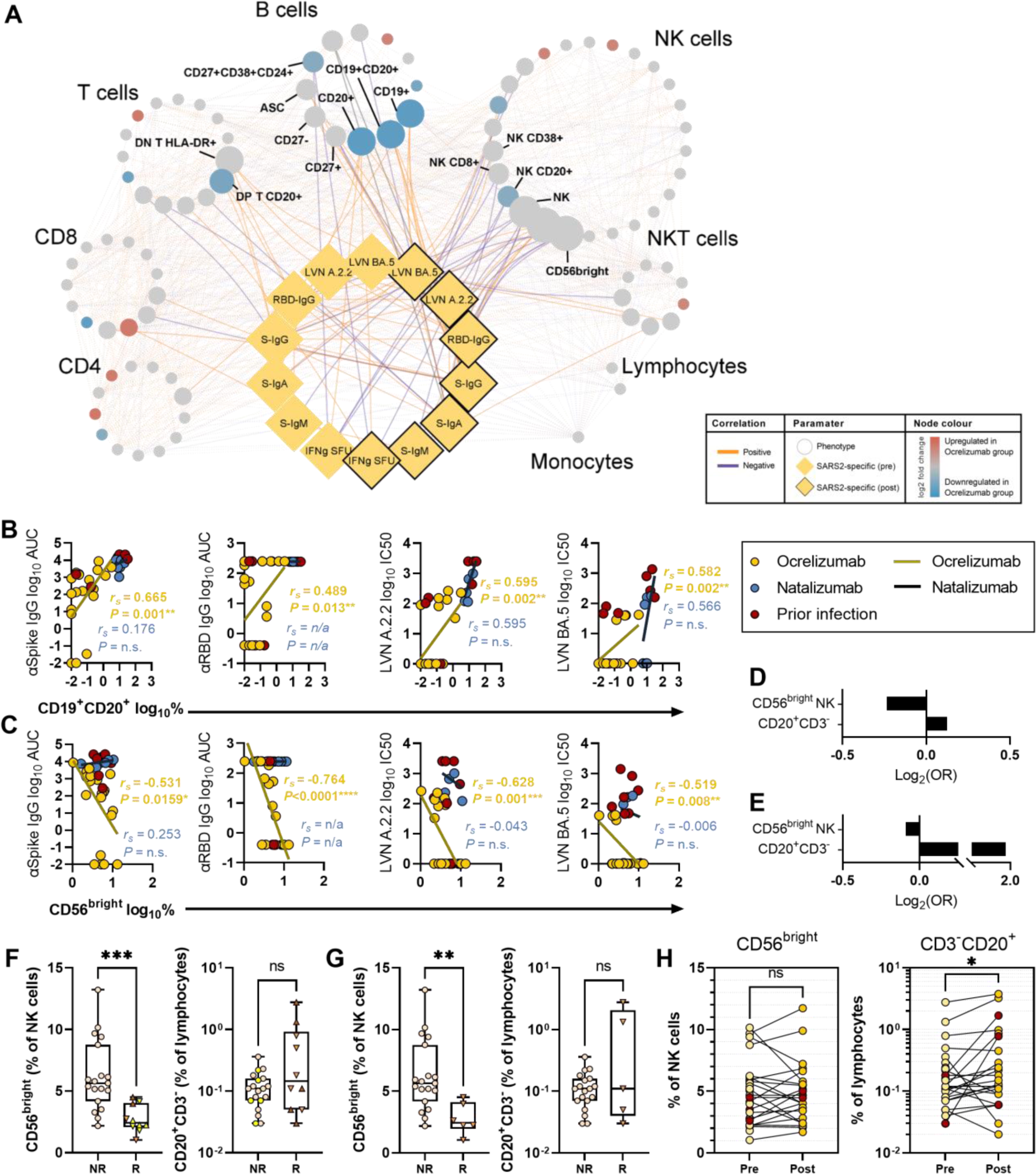
Association of immune phenotype with vaccine response in pwMS receiving ocrelizumab. **(A)** Spearman’s correlation network describing the relationship between immune phenotype pre-vaccination and vaccine response parameters in pwMS receiving ocrelizumab. Nodes are manually clustered into related immune cell lineages. Node colour denotes differences from the natalizumab group (from Figure 2B). Node size corresponds to the number of edges formed with vaccine response measures. Positive correlations are shown in orange and negative correlations are shown in purple. Phenotype-to-response edges are bold, while phenotype-to-phenotype edges are faded. Vaccine response measures (yellow diamond nodes) are shown pre-third dose (no outline) and post-third dose (black outline). **(B-C)** Relationship of pre-vaccination CD19^+^CD20^+^ B cells (% of lymphocytes) (B) and CD56^bright^CD16^−^ NK cells (% of all NK cells) (C) with antibody response to vaccination, independently assessed by Spearman’s correlation analysis. **(D-E)** LASSO logistic regression models for prediction of effective serological neutralisation of ancestral SARS-CoV-2 were constructed for the full ocrelizumab cohort (D) and independently for pre-vaccination non-responders (E) from the original 101 phenotype parameters. **(F-G)** Comparison of pre-vaccination selected parameters between pwMS on ocrelizumab who did (‘R’) and did not (‘NR’) achieve effective ancestral neutralisation following third vaccine dose. **(H)** Change in frequency of selected immune cell populations across pre to post third vaccination time points.

Unexpectedly, natural killer (NK) cell populations were associated with vaccine response in ocrelizumab treated patients (Figure 3C). CD16^−^CD56^bright^ NK cells at the time of vaccination (both as a proportion of NK cells and as a proportion of lymphocytes) were strongly negatively correlated with anti-Spike and anti-RBD IgG titres post-vaccination, and with neutralisation of ancestral and omicron BA.5 variants (Figure 2C). This association was not observed in the natalizumab group (Figure 3C).

In a penalised multivariate logistic regression model, CD3^−^CD20^+^ B cells (% of lymphocytes) and CD56^bright^ NK cells (% of total NK cells) were selected as predictors of effective neutralisation of ancestral SARS-CoV-2 by pwMS receiving ocrelizumab with an area under the receiver operator characteristic (AUROC) curve of 0.91 (CD56^bright^ NK cells: OR = 0.85; CD3^−^CD20^+^ B cells: OR = 1.09; Figure 3D). The same parameters were selected when regularisation was performed for only the individuals who were non-responders (A.2.2 LVN < 20) prior to the third dose (AUROC = 0.98; CD56^bright^ NK cells: OR = 0.94; CD3^−^CD20^+^ B cells: OR = 3.92; Figure 3E). All participants with pre-vaccination CD3^−^CD20^+^ B cell frequencies ≥0.5% of lymphocytes achieved effective neutralisation of ancestral virus, and frequencies of CD56^bright^ NK cells were significantly lower in responders than non-responders (Figure 3F-G). While CD3^−^CD20^+^ B cell frequency changed over time, consistent with gradual reconstitution of the B cell compartment, CD56^bright^ NK cell frequency was stable across the pre to post vaccination time points (Figure 3H).

## DISCUSSION

PwMS receiving B cell-depleting therapies are among the most vulnerable to COVID-19 and other vaccine-preventable infections. With updated prophylactic antiviral measures becoming available, we sought to evaluate the level of protection afforded this group by booster vaccination, and to identify biomarkers and modifiable determinants of protective immunity to inform future vaccination strategies. Consistent with previous studies,^10^ approximately half of participants were seropositive for SARS-CoV-2 RBD-IgG following a third vaccine dose. The quality of the antibody response in seropositive individuals was largely inadequate: assessed against target viral neutralisation values for protection from infection, 25% of the ocrelizumab cohort demonstrated effective neutralisation of ancestral SARS-CoV-2, and 14% against the immune evasive omicron BA.5 variant. Thus, pwMS on B cell-depleting therapy remain inadequately protected from COVID-19 with three vaccine doses, and are amongst the poorest protected populations.

The major approach to improving vaccine immunity in immunocompromised groups has been repeated booster dosing with homologous or variant vaccines. In our cohort, booster vaccination improved the median IgG titre and the frequency of virus-specific IFNγ-secreting T cells, however less than one-third of seronegative participants seroconverted with a third vaccine dose, and only one participant who was seronegative prior to the third dose achieved effective viral neutralisation. Although limited, data on response to a fourth vaccine dose in patient groups receiving B cell depleting therapy suggest that an additional 24-38% of seronegative patients may be expected to seroconvert with a fourth dose ^20, 21^. It is worth noting that these studies and others have relied on detection of serum NC-IgG to identify participants with prior SARS-CoV-2 infection. This criteria is flawed as the four participants in our ocrelizumab cohort with PCR-confirmed SARS-CoV-2 did not have detectable NC-IgG.

Time since infusion has been suggested as a key modifiable determinant of vaccine response, with vaccine administration at the end of an infusion cycle recommended. The standard dosing interval for ocrelizumab is six months and some centres have implemented delayed infusions to maximise B cell reconstitution prior to vaccination. While a relationship between time since ocrelizumab infusion and antibody response has been reported,^22, 23^ it is participants that have been vaccinated more than six months post-infusion that appear to drive this association. A similar study to ours in which the majority of participants are vaccinated *within* six months of infusion did not find an association with vaccine response.^13^ Given the risks associated with delaying treatment, the lack of a clear benefit observed in our study, and the low rates of protection against prevalent variants (14.3% against omicron BA.5 in our study), indiscriminate prophylaxis for pwMS on B cell depletion may be appropriate where possible.

The problem of inadequate vaccine response in this cohort extends beyond COVID-19, and raises the potential for personalised approaches to vaccination. Several studies have suggested B cell reconstitution as a biomarker of response, with 40 B cells /uL ^13^ (approximately 1.6% of lymphocytes) suggested as a pre-vaccination target in pwMS, and 10 B cells /uL (approximately 0.4% of lymphocytes) suggested in a study of rheumatoid arthritis patients receiving B cell depletion,^24^ although three quarters of participants were >6 months post infusion. This value is supported by the present study in which all four participants with CD3^−^ CD20^+^ B cell frequencies greater than 0.4% of lymphocytes achieved seroconversion and effective serum neutralisation of ancestral SARS-CoV-2 virus.

The majority of responders in our study were able to achieve effective viral neutralisation despite having low pre-vaccination B cell frequencies. Comprehensive evaluation of multiple immune phenotypes identified a novel predictor of antibody response in CD56^bright^ NK cells, which inversely correlated with antibody titres and neutralisation, and, in conjunction with B cell reconstitution in a multiple logistic regression model, considerably improved stratification of responders. CD56^bright^ NK cells are an immunomodulatory population of NK cells, which are cytotoxic toward proliferating and autoreactive CD4^+^ T cells *in vitro.*^25–29^ In MS, they have been reported to be elevated in the cerebrospinal fluid (CSF) and present in periventricular lesion of the brain, although their role is unclear.^30, 31^ Several DMTs (not including ocrelizumab and natalizumab) have been reported to alter the frequency of CD56^bright^ NK cells in the periphery,^32–45^ and an increase (or lack of decrease) in CD56^bright^ NK cells has been associated with response to treatment with IFNβ, dimethyl fumarate, fingolimod and daclizumab,^34, 35, 46–48^ suggesting a potential immunoregulatory role in MS. In this study, CD56^bright^ NK cell frequency was similar between ocrelizumab and natalizumab treatment groups, however was only associated with poor antibody response to vaccination in the ocrelizumab cohort. A potential explanation, therefore, is that the negative influence of CD56^bright^ NK cells on the antibody responses is imperceptible under normal circumstances but is unmasked in the absence of B cells. The relationship between CD56^bright^ NK cells and antibody responses to vaccination should be validated in an independent prospective study, as it may represent a novel biomarker of vaccine response and shed light on the mechanistic relationship between CD56^bright^ NK cells and MS disease pathogenesis.

Collectively, we highlight severe immune impairment in pwMS on B cell-depleting therapy, and provide insights into the changes in immune phenotype associated with ocrelizumab use relevant to disease pathogenesis and how the immune phenotype relates to aspects of cellular and humoral response to vaccination. The data presented here support the use of prophylactic measures for protection against COVID-19 in pwMS receiving B cell depleting therapies, and support the potential of immune phenotype markers for the personalisation of vaccination strategies in this highly vulnerable group.

---

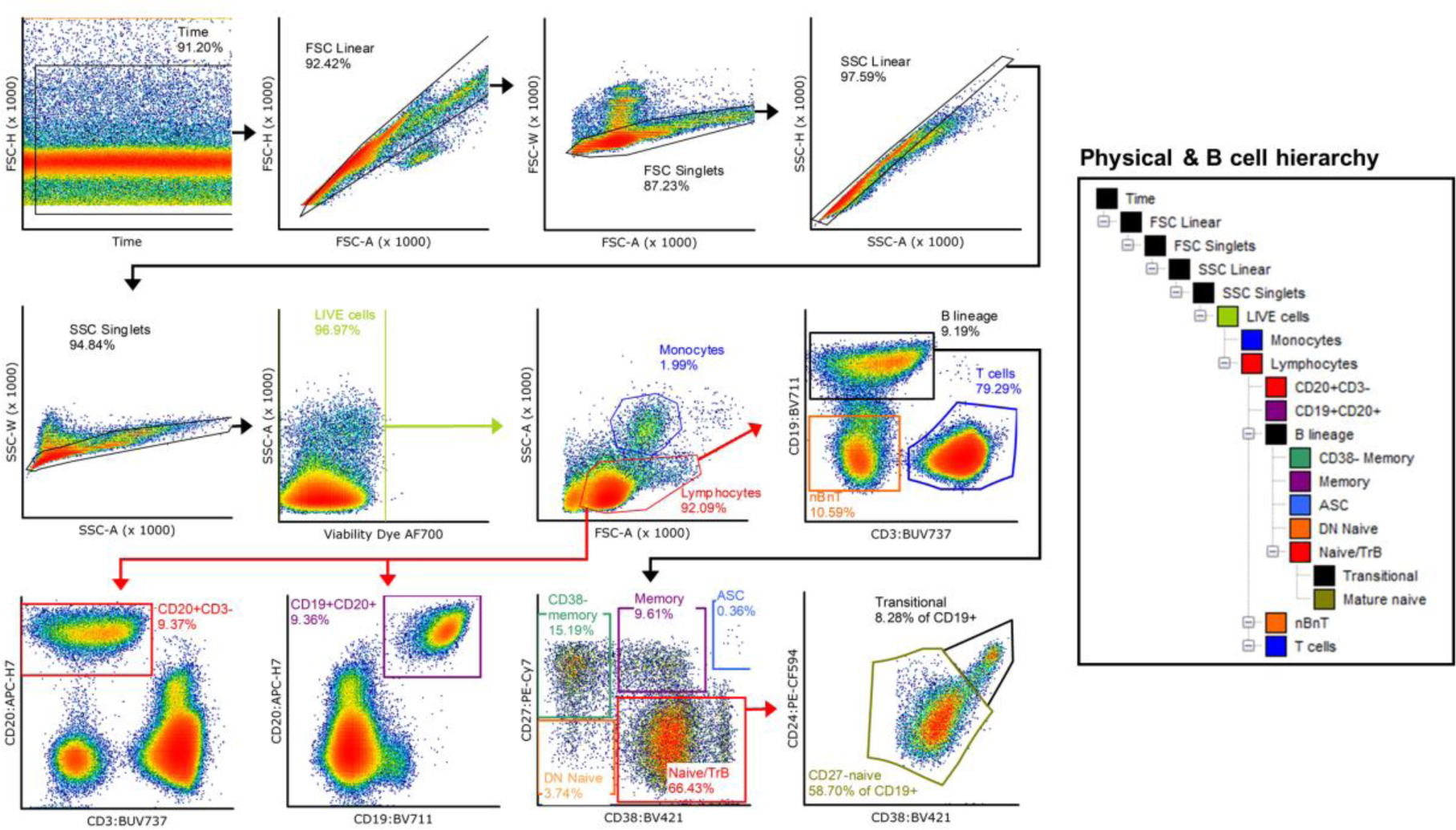
Supplementary Figure 1

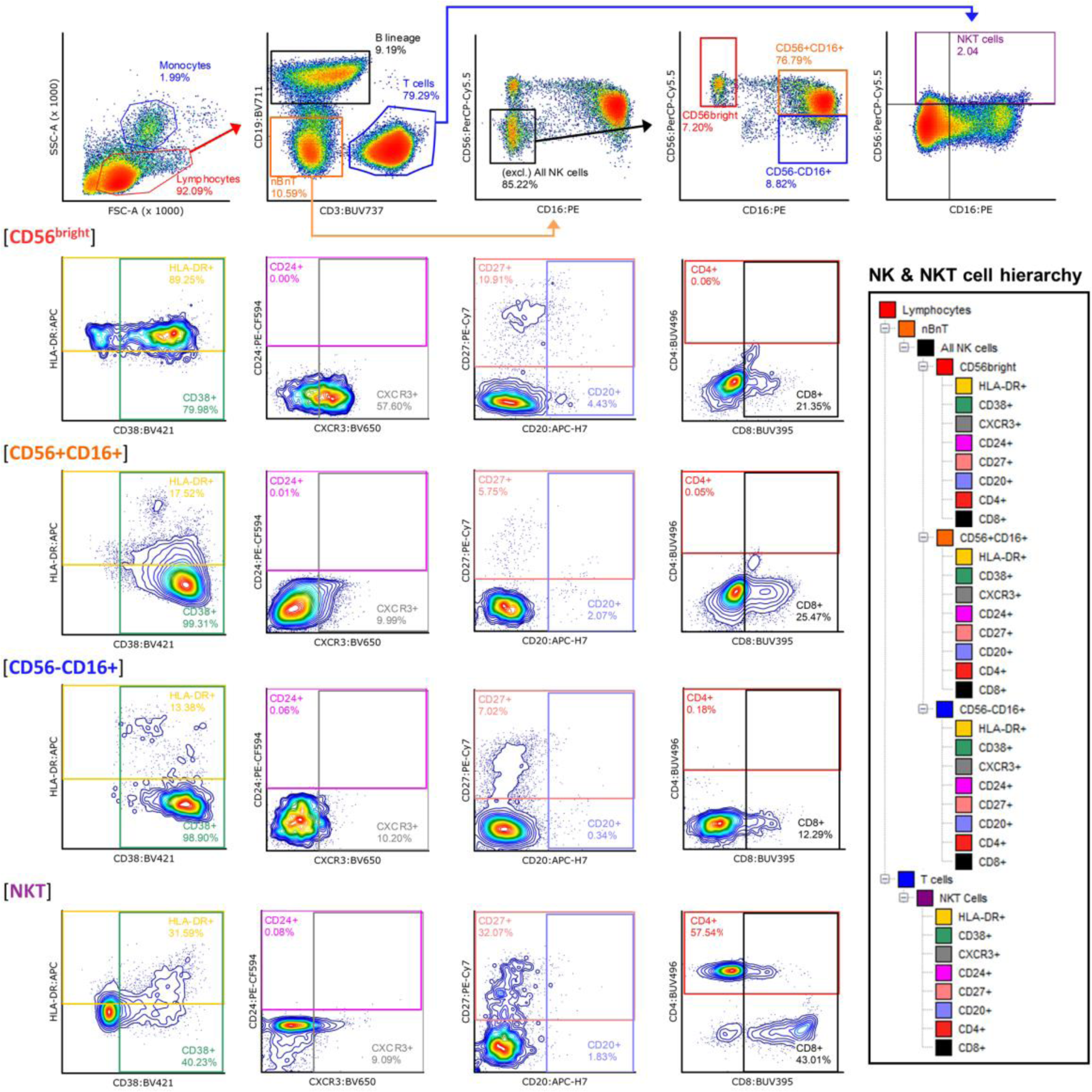
Supplementary Figure 2

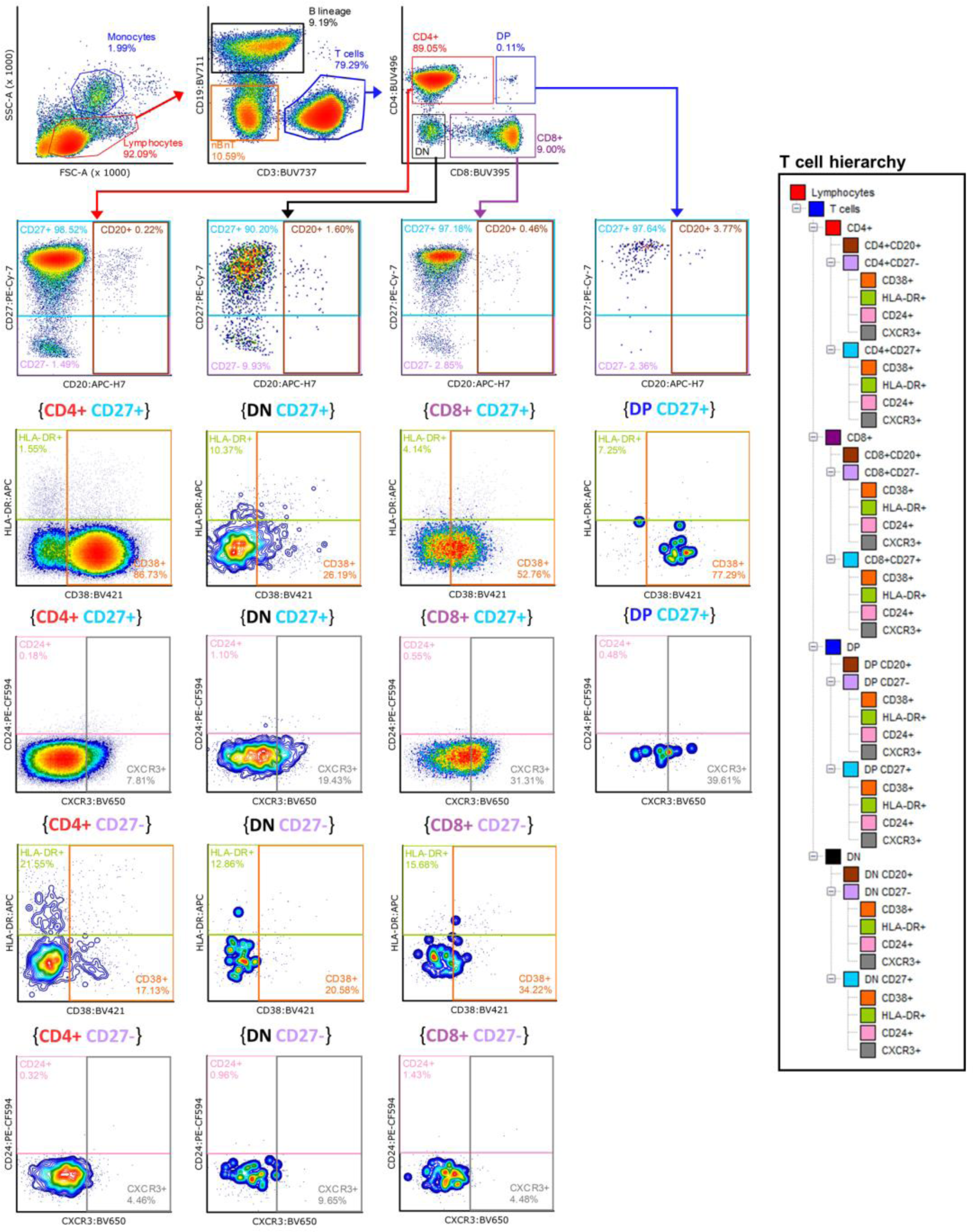
Supplementary Figure 3

## METHODS

### Standard Protocol Approvals, Registrations, and Participant Consents

The study was approved by the Central Adelaide Local Health Network Human Research Ethics Committee (Approval Number 15083), and conducted from November 2021 – August 2022 (ACTRN12623001249640). Study participants were recruited from the Royal Adelaide Hospital Specialty Vaccination Clinic and informed consent was provided by all participants. Inclusion criteria were ability to understand requirements of the study, provide written informed consent and attend follow-up; age 18+ years; diagnosis of RRMS, PPMS, SPMS; receiving ocrelizumab or natalizumab as primary treatment with the last dose received within 24 months of vaccine administration; have elected to receive a third COVID-19 vaccine dose with a Therapeutic Goods Administration (TGA) approved mRNA-platform vaccine. Past SARS-CoV-2 infection was determined by record of positive nasopharyngeal swab by PCR and confirmed via verbal history taken from all participants. All participants were followed-up by telephone February 2023 (approximately 6 months post-final study visit), and incidence and severity of COVID-19 (hospitalisation and treatment required) was self-reported at this time, and confirmed with electronic medical records.

### SARS-CoV-2 Spike protein production and ELISA

Anti-SARS-CoV-2 Spike IgM, IgA and IgG were quantified by in-house ELISA, as previously described.^49, 50^ Plates were coated with prefusion SARS-CoV-2 Spike ectodomain (isolate WHU1, residues 1-1208) with HexaPro mutations (kindly provided by Dr Adam Wheatley),^49^ and participant sera serially diluted and end point titres calculated and expressed as area under the curve (AUC). AUC calculations were performed using Prism v9.0.0 (GraphPad Software Inc.).

Anti-SARS-CoV-2 Spike Receptor-Binding Domain (RBD) and Nucleocapsid (NC) IgG titres were quantified using the ‘Elecsys Anti-SARS-CoV-2 S’ and ‘Elecsys Anti-SARS-CoV-2’ assays on the Cobas system (Roche), through the state pathology service, SA Pathology. The quantitation range for detection of anti-RBD IgG in this assay is 0.4-250 U/mL. This assay is widely available through pathology services globally and can be compared with other assay by conversion to WHO Standard International Units, where 1 U/mL is comparable to ∼1.029 BAU/mL.^51^

### SARS-CoV-2 live-virus neutralisation assay

Rapid high-content neutralisation assay with HEK-ACE2/TMPRSS cells, HAT-24, cells was performed as previously described.^52, 53^ HAT-24 cells were seeded in 384-well plates at 16 × 10^3^ cells/well in the presence of the live cell nuclear stain Hoechst-33342 dye (NucBlue, Invitrogen) at a concentration of 5% v/v. Two-fold dilutions of patient serum samples were mixed with an equal volume of SARS-CoV-2 virus solution standardized at 2xVE_50_ and incubated at 37°C for 1 h before adding 40 μL, in duplicate, to the cells. Viral variants used were the omicron sub-variant BA.5, and the ‘ancestral’ virus (A.2.2) from clade A and presenting no amino acid mutations in Spike (similar to Wuhan ancestral variant and vaccine strain). Plates were incubated for 24 h post infection and entire wells were imaged by high-content fluorescence microscopy, cell counts obtained with automated image analysis software, and the percentage of virus neutralisation was calculated with the formula: %N = (D-(1-Q)) × 100/D, as previously described.^52, 53^ Sigmoidal dose-response curves and IC_50_ values (reciprocal dilution at which 50% neutralisation is achieved) were calculated with Prism v9.0.0 (GraphPad Software Inc.).

The threshold for effective neutralisation was derived from a published value of 20.2% of the mean neutralisation titre (IC_50_) of first-wave convalescent individuals.^14^ This value correlated with 50% protection from real-world infection early in the COVID-19 pandemic. In our study, 20 convalescent sera from an early wave in South Australia were included in the same experimental run as the study samples, and 20.2% of the mean IC_50_ for this group calculated to be 14.7 (95% CI 10.5 to 20.7). As the lowest dilution factor tested for all samples was 20, and this was within the 95% CI calculated by Khoury *et al*, effective neutralisation of both the ancestral and omicron BA.5 variants was defined as an IC_50_ ≥ 20. While the present study extrapolates the same value for ‘effective neutralisation’ to the analysis of omicron BA.5, an equivalent correlate for the omicron BA.5 variant has not been published.

### IFNγ ELISpot

IFNγ ELISpots were performed in-house and counts are comparable with previous reports from our group.^54–56^ Briefly, multiscreen-IP HTS plates (Merck Millipore) were coated with anti-human IFN-γ (clone 2G1, Thermo Fisher). PBMCs were thawed by dropwise addition of complete media (20% FCS) with Benzonase® nuclease (Merck Millipore) to prevent aggregation, and stimulated for 18h with 4 pools of overlapping peptides spanning the entire length of the spike glycoprotein of the USA-WA1/2020 strain. Individual peptides are 17- or 13-mers, with 10 amino acid overlaps, obtained through BEI Resources, NIAID, NIH (*Peptide Array, SARS-Related Coronavirus 2 Spike (S) Glycoprotein, NR-52402*). Secreted IFN-γ was detected with anti-human IFN-γ:biotin (Clone B133.5; ThermoFisher) followed by streptavidin:HRP (BD Biosciences) and AEC substrate (BD Biosciences). Developed spots were counted automatically by use of an ELISpot reader (Cellular Technology Ltd., Bonn, Germany), background (untreated well counts) subtracted from stimulated wells, and counts presented as the sum of the four pools per 10^6^ cells.

### Immune phenotyping

Immune phenotyping was performed using a previously described and validated panel/protocol.^49^ Briefly, thawed PBMCs were stained in a U-bottom plate with 30 μL antibody master-mix for 20 min in the dark. Stained PBMCs were washed twice with 200 μL FACS wash, centrifuged at 300×g for 5 min and fixed with 200 μL FACS Fix for 20 min, at RT in the dark. Fixed cells were then centrifuged 300×g for 5 min, washed in 200 μL FACS wash then spun 300×g for 5 min, and resuspended in 50 μL FACS wash for analysis using the BD FACS Symphony within 24 hours.

Staining and acquisition was performed in three batches. To control for batch effects, the BD FACS symphony lasers were calibrated with dye conjugated standards (Cytometer Set &Track beads) before each run. All samples were acquired with all 28 PMTs recording events. All PMT voltages were adjusted to unstained negative control baseline, typically log scale 10^2^. Antibodies were titrated for optimal signal-over-background so that single positive stains sat within log scale 10^3^–10^5^ of designated PMT. Compensation was set with beads matched to each panel antibody combination using spectral compensation using FlowJo Software v10 (BD Biosciences). Exported FCS files had compensation values adjusted manually post-acquisition on a file-by-file basis in FCS Express v6 (De Novo Software) and gates manually adjusted between batches as required. Once compensated, low data quality events were excluded based upon time acquired (at the sample acquisition start and before sample exhaustion), with further time exclusion gates based on blockages or unexplained loss of events for a period of time during acquisition. Events positive for LIVE/DEAD staining were removed, and events were gated for FSC-H/-A as well as SSC-H/-A linearity, and restricted FSC-W and SSC-W values for doublet discrimination. Live single cells were then broadcast on a SSC-A/FSC-A plot to determine size and complexity. Lymphocytes and monocytes were expressed as a percentage of viable single cells (based on physical gates), and subpopulations expressed as a percentage of parent gate, unless otherwise indicated.

### Statistics and Data Visualisation

All statistical analyses were performed using Prism v9.0.0 (Graphpad Software Inc.), Stata Statistical Software: Release 14.2 (StataCorp), or R v4.3.1. All tests were two-tailed and no assumptions were made about the distribution of the data sets; non-parametric tests were used in all cases for comparisons. Accordingly, Mann-Whitney and Kruskal-Wallis tests with Dunn’s correction were applied to pair-wise and multiple comparisons, respectively.

#### Exploratory Comparisons

Differences in pre-vaccination immune phenotype parameters between treatment groups were identified by Multiple Mann-Whitney tests, with Two-stage step-up method employed to correct for multiple comparisons. Statistical significance was determined as a false discovery rate (FDR) < 0.01.

#### Regression Modelling

Multiple linear regression by least squares modelling was applied to post-vaccination antibody titres (S-IgM, S-IgA, S-IgG, RBD-IgG), neutralisation data (A.2.2 and omicron BA.5) and ELISpots, as well as change in ELISpot counts pre to post vaccination for the ocrelizumab cohort. Assumptions of normality of residuals was tested using D’Agostino-Pearson omnibus (K2) and of linearity using visual inspection of scatter plots. Variables included were age, sex (M/F), time between infusion and vaccination, primary course vaccine type (BNT162b2/ChAdOx1), and time between second and third dose. Association of pre-existing immunity (i.e. pre-vaccination antibody and neutralisation titres and ELISpot counts) with change in ELISpot count pre to post vaccination was assessed in a separate multiple linear regression analysis. Multiple linear regression modelling was conducted using Prism v9.0.0 (GraphPad Software Inc.). Association between immune phenotype parameters and vaccine response was assessed by multiple logistic regression for a binary outcome of effective neutralisation of SARS-CoV-2 A.2.2 (IC_50_ ≥ 20). Covariate selection was achieved with penalised LASSO regularisation for logistic regression using the Glmnet package (v4.1-7) in R, which reduces coefficients of multi-collinear and non-significant variables to zero. This approach resulted in a multiple logistic regression model for CD20^+^ B cell and CD56^bright^ NK cell frequencies, with a conservative (penalised) odds ratio estimate. A limitation of the regularisation approach is that a meaningful p value and confidence interval cannot be estimated, and the sample size of the study was not sufficiently large to allow bootstrapping.

#### Dimensionality Reduction

For visual comparison of B cell phenotype between treatment groups, phenotype data (FCS files) for 13 ocrelizumab and 13 natalizumab samples acquired in the same batch were concatenated. CD19^+^ events from natalizumab samples were down-sampled such that the analysis included an equal number of B cell events from each treatment group. Dimensionality reduction parameters by t-distributed stochastic neighbor embedding (tSNE) were estimated based on 100,000 events (Perplexity: 90; Iterations: 1500) for major B cell parameters (CD24, CD38, CD27) using FCS Express v6 (De Novo Software), and five FlowSOM clusters defined using FlowJo Software v10 (BD Biosciences). Concatenated data from the representative samples were displayed as a coloured dot plot, and the relative frequencies of B cell subsets for each treatment group compared by side-by-side density plots (of equal event count) and pie-charts. As a representative figure, no statistical analysis was applied.

#### Pearson’s Correlation Analysis

Spearman’s correlations were performed using Prism v9.0.0 (GraphPad Software Inc.). Relationships between pre-vaccination immune phenotype variables, vaccine response measures, and changes in immune phenotype associated with treatment in the ocrelizumab group were visualised using Cytoscape v3.9.1. Network edges between immune phenotype parameters and vaccine response measures were constructed for significant (p < 0.05) Spearman’s rank correlation coefficients (*r_S_*) of less than −0.3 and greater than 0.3, and immune phenotype nodes sized and coloured to reflect relatedness (number of shared edges/correlations) with vaccine response measures and significant differences to the natalizumab cohort, respectively.

## Data Availability

Phenotype and correlation data are available as a network (SIF) file, and raw values can be obtained by contacting G.B.P.

## Acknowledgments

We thank everyone who contributed to this study, particularly those who participated in the trials during the height of the pandemic. We would also like to thank the staff of the Royal Adelaide Hospital Medical Day Unit and Specialist Vaccination Clinic, and acknowledge Dennis Penglis and Tina Petrou for their assistance with serological studies.

## Author contributions

Conceptualisation: GBP, JR, PH.

Methodology: GBP, MJT, CSC, CMH, ST, BG-B.

Formal analysis: GBP, CMH, MJT, SS, AELY.

Investigation: GBP, CMH, CSC, MJT, AELY, SK, JDZ, AAg, VM, AAk.

Visualization: GBP.

Funding acquisition: GBP, SCB, BGB, PTC, PH.

Project administration: GBP, KW, JY, WW, MBR, PH.

Supervision: PRH, CMH, MGM, PH, SCB, BGB, PTC.

Writing – original draft: GBP.

Writing – review & editing: All authors reviewed the manuscript.

